# Effect of pregnancy intention at conception on the continuity of care in maternal healthcare services use in Somalia: Evidence from first national health and demographic survey

**DOI:** 10.1101/2024.04.03.24305262

**Authors:** Md Badsha Alam, Shimlin Jahan Khanam, Md Awal Kabir, Ibrahim Yasin Khalif, Md Nuruzzaman Khan

## Abstract

**Background:** Unintended pregnancies pose a significant challenge to maternal healthcare service utilization and continuity of care (CoC) in low-resource settings. This study investigates the impact of pregnancy intention at conception on CoC in maternal healthcare service use in Somalia.

**Methods:** Data comprising 7,079 mothers were extracted from the First National Health and Demographic Survey of Somalia conducted in 2020, with the condition that they had given birth within the three years preceding the survey. Level of Continuity of Care (CoC), categorized as lowest, middle, or highest, in receiving maternal healthcare services, was considered as the explanatory variable and assessed based on the receipt of antenatal healthcare (ANC, <4, ≥4), skilled birth attendance (SBA), and postnatal healthcare (PNC, within 48 hours of birth). Mother’s intention of most recent pregnancy at conception was the primary exposure variable. Unadjusted and adjusted multilevel multinomial logistic regression models were used to assess the effect of unintended pregnancy on the level of CoC completion.

**Results:** Approximately 38% of live births in Somalia were unintended at conception. Only 2.4% of mothers achieved full CoC, with higher rates observed among those with intended pregnancies (3.0%) compared to those with mistimed (1.6%) or unwanted pregnancies (1.1%). Mothers having mistimed (aOR, 0.59, 95% CI, 0.37-0.95) and unwanted (aOR, 0.28, 95% CI, 0.10-0.79) pregnancies had significantly lower odds of achieving moderate and higher levels of CoC compared to those having intended pregnancies, even after adjusting for socio-demographic factors.

**Conclusion:** Unintended pregnancies are associated with lower level of CoC in maternal healthcare service use in Somalia. Strengthening family planning services, promoting contraceptive use, enhancing sexual education, and raising awareness about the importance of maternal healthcare services are essential steps to reduce unintended pregnancies and improve maternal and child health outcomes in the country.

## Introduction

Somalia grapples with a formidable challenge in maternal and child mortality, marked by an estimated maternal mortality rate of 692 deaths per 100,000 live births, three times higher than the global average (223 per 100,000 live births) and twice that of low- and middle-income countries (LMICs, 368 per 100,000) (1, 2). This places Somalia among the countries with very high maternal mortality rates, alongside Yemen and South Sudan (3, 4). Child mortality rates also remain distressingly high, with 68 infant deaths and 28 stillbirths per 1,000 births, approximately double the average infant mortality rate of 43 per 1,000 births in LMICs (5-7). These stark statistics underscore Somalia’s formidable challenge in attaining Sustainable Development Goals related to maternal (Goal 3.1: reducing the maternal mortality rate to 70 per 100,000 live births) and child (Goal 3.2: reducing neonatal and infant mortality to 12 and 25 per 1,000 live births, respectively) (8). They also illuminate the nation’s struggle to keep pace with global advancements in maternal and child health.

The higher rates of maternal and child mortality in Somalia reflect broader challenges related to maternal healthcare access, infrastructure, and socioeconomic disparities (1, 9, 10). Limited access to maternal healthcare services is a significant concern, with recent data indicating that only 23% of Somali mothers receive at least four ANC consultations, a significant drop from the 85% who receive at least one ANC (1, 11). Additionally, only around half of deliveries occur in hospitals, with a mere 10% receiving PNC (1, 11). These statistics reveal that a majority of mothers in Somalia lack access to maternal healthcare services, and among those who do, there is inadequate continuity of care (CoC), increasing vulnerability to complications for both mothers and newborns. Furthermore, lower rates of exclusive breastfeeding, coupled with the prevalence of communicable diseases and malnutrition, pose additional challenges to child health and development in Somalia (1). Addressing these issues necessitates ensuring CoC in receiving maternal healthcare services.

The high prevalence of unintended pregnancy poses a significant concern for Somalia, similar to other LMICs (12, 13). Recent estimates suggest that around 40% of pregnancies resulting in live births in Somalia are unintended at conception (1). Restricted access to family planning and safe abortion services are key factors contributing to this issue (14). However, these circumstances create two potential barriers to accessing maternal healthcare services with continuity: (i) missed family planning visits represent lost opportunities for healthcare providers to counsel mothers on accessing maternal healthcare services, and (ii) adverse effects of unintended pregnancy, such as depression and negative feelings about the index pregnancy, restrict mothers from accessing maternal healthcare services (15). Additionally, unintended pregnancies are often identified late, delaying the initiation of ANC and negatively impacting the achievement of CoC (15, 16). Despite these challenges and indication of unintended pregnancy on CoC, there is a lack of understanding regarding the effects of unintended pregnancy on CoC in maternal healthcare services in Somalia, as no previous study has explored this issue (15). Therefore, this study aims to investigate the impact of unintended pregnancy on CoC in accessing maternal health services in Somalia.

## Methods

### Data Source and Sampling Methodology

This study utilized data from the Somali Health and Demographic Survey (SHDS) 2020, the first nationally representative survey conducted by the Somalian Government, following the sampling process of the Demographic and Health Survey programs of the USA. The survey employed a stratified multi-stage cluster sampling design, including a three-stage sample selection procedure in urban and rural areas and a two-stage process in nomadic areas, where nomadic people migrate and temporarily reside. Initially, 55 sampling strata were selected across the country, with 47 included in the survey due to security concerns. A total of 1,433 enumeration areas (EAs) were chosen from the selected 47 strata, comprising 770 EAs from urban areas, 488 from rural areas, and 175 from nomadic areas. In the subsequent stages, 545 EAs were selected, with 220 allocated to urban strata, 150 to rural strata, and 175 to nomadic areas. The third stage involved selecting 30 households from each EA, resulting in 16,360 households for interview. Out of these households, 18,202 eligible women were identified based on inclusion criteria, with 16,715 successfully interviewed, yielding a response rate of 91.8%. Interested readers can refer to the SHDS report for detailed information on the survey and sampling procedure (1).

### Analyzed Sample

We analyzed a subset of 7,079 mothers extracted from the total of 16,715 mothers included in the survey. Inclusion criteria were: (i) having given birth to at least one live child within three years prior to the survey date, (ii) responding to questions regarding maternal healthcare services use during pregnancy (focusing on the most recent pregnancy if multiple pregnancies occurred), and (iii) reporting their intention at conception of this pregnancy.

### Explanatory Variable

Level of Continuity of Care (CoC) served as the explanatory variable, derived from women’s responses regarding the utilization or non-utilization of ANC, SBA, and PNC. Services received from medically trained providers were considered all cases. CoC levels were categorized as highest (four or more ANC visits, assisted birth by SBA, and PNC within 48 hours of birth), middle (at least two of the following: four or more ANC visits, SBA, and PNC services), and lowest (no SBA, no PNC, and less than four ANC visits). These classifications followed the WHO guidelines for maternal healthcare services utilization and provider definitions. According to WHO guidelines, during pregnancy, each mother should receive at least four skilled ANC visits, be assisted by SBA during delivery, and receive PNC (for both women and newborns) within 48 hours of birth from skilled healthcare personnel. The SDHS collected information on maternal healthcare services utilization and providers through various questions related to ANC, SBA, and PNC.

### Exposure Variables

The primary exposure variable was mother’s intention at conception of their most recent pregnancy resulting in a live birth. Eligible mothers were inquired about their pregnancy intentions concerning their last child born within three years of the survey date. They were asked, “*Did you intend to become pregnant at that time*?” with response options of “Yes” or “No”. Those who answered “No” were further questioned, “Did you plan to have a child later or did you not desire any more children?” Responses were then categorized as “wanted” (if they responded “yes” to the initial question), “mistimed” (if they answered “later” to the subsequent inquiry), and “unwanted” (if they indicated “no more” in the follow-up question). The reliability and validity of this data collection and categorization method have been consistently demonstrated across various regions, including Asian and African countries (11, 15, 17, 18).

### Covariates

The association between CoC level and maternal pregnancy intention at conception for the index child was adjusted for various factors at the individual, household, and community levels. These factors were identified through a literature review and forward regression analysis (13, 16, 19-21). Individual-level factors included the mother’s age at childbirth, education level, employment status, decision-making autonomy, and parity. Household-level factors consisted of an index of mass media exposure and household wealth quintile. The mass media exposure index was derived from respondents’ self-reported weekly exposure to newspapers, radio, and television. The household wealth quintile variable was generated by the survey authority using principal component analysis, as presented in the survey data (1). Community-level factors encompassed the respondent’s place of residence and administrative region.

### Statistical Analysis

Descriptive statistics were utilized to describe the characteristics of the respondents. Various combinations of maternal healthcare services across the CoC were examined, both overall and stratified by intention at conception. Dropout rates across CoC levels to PNC were computed. Unadjusted and adjusted multilevel multinomial logistic regression models were utilized to evaluate the association between pregnancy intention and CoC level. Sampling weights were incorporated, and outcomes were presented as odds ratios (OR) with corresponding 95% confidence intervals (CI). Stata software version 17.0 was employed for all analyses.

### Ethics Approval and Consent to Participate

The survey analyzed was approved by the institutional review board of SHDS, and informed consent was obtained from all participants. All necessary patient/participant consent has been obtained, and the appropriate institutional forms have been archived. Permission to access the survey and conduct this research was obtained, and all methods were performed in accordance with relevant guidelines and regulations. No separate ethical approval was required to conduct this study.

## Results

### Background characteristics of the respondents

Table 1 presents the background characteristics of the respondents. Among all pregnancies, 62.5% were intended at conception, while 27.6% were mistimed, and 9.9% were unwanted. The majority of mothers fell within the 20-34 years age group (71.9%), with smaller proportions in younger and older age categories. Approximately 83% of the total mothers were illiterate, followed by those with primary (12.5%) and secondary education (4.6%). A notable percentage of mothers were not employed (94.0%), and about half were involved in decision-making regarding their healthcare (54.3%). A considerable proportion had no exposure to mass media (82.7%). Regarding residence, a significant portion of respondents lived in urban areas (35.0%), while others resided in rural (30.3%) or nomadic (34.8%) settings.

**Table 1.** Background characteristics of the respondents, SHDS, 2019 (N=7,079)

| Characteristics | Percentages | 95% CI |
| --- | --- | --- |
| Exposure variable: pregnancy intention at conception |  |  |
| Wanted | 62.5 | 60.2-64.7 |
| Mistimed | 27.6 | 25.9-29.4 |
| Unwanted | 9.9 | 8.8-11.1 |
| Individual level factors |  |  |
| Mother's age (in years) |  |  |
| ≤19 | 7.4 | 6.7-8.2 |
| 20-34 | 71.9 | 70.4-73.3 |
| ≥35 | 20.7 | 19.4-22.1 |
| Mother's education |  |  |
| No education | 82.9 | 81.1-84.5 |
| Primary | 12.5 | 11.2-14.0 |
| Secondary | 4.6 | 3.9-5.4 |
| Higher |  |  |
| Mother's employment status |  |  |
| Not employed | 94.0 | 93.2-94.8 |
| Employed | 6.0 | 5.2-6.8 |
| Mother's decision-making autonomy |  |  |
| Mothers involved | 54.3 | 51.6-57.0 |
| Mothers not involved | 45.7 | 53.1-48.4 |
| Parity |  |  |
| ≤2 | 27.6 | 26.2-29.1 |
| 3-4 | 29.4 | 27.9-31.0 |
| >4 | 43.0 | 41.2-44.8 |
| Household level factors |  |  |
| Mass media exposure |  |  |
| Not exposed | 82.7 | 81.0-84.3 |
| Moderately exposed | 14.6 | 13.2-16.0 |
| Highly exposed | 2.7 | 2.1-3.5 |
| Wealth quintile |  |  |
| Lowest | 23.7 | 20.7-27.0 |
| Second | 20.8 | 18.7-23.2 |
| Middle | 18.5 | 15.9-21.5 |
| Fourth | 20.4 | 18.1-22.8 |
| Highest | 16.6 | 14.5-18.9 |
| Community level factors |  |  |
| Place of residence |  |  |
| Urban | 35.0 | 30.9-39.2 |
| Rural | 30.3 | 24.0-37.3 |
| Nomadic | 34.8 | 30.8-39.0 |
| Administrative region |  |  |
| Northwest | 35.3 | 30.6-40.3 |
| Northeast | 30.0 | 24.7-35.8 |
| Central | 26.9 | 23.0-31.3 |
| South | 7.8 | 5.7-10.6 |

### Integration of Independent Maternal Healthcare Service Indicators into a Continuum of Care

S1-Table 1 presents the distribution of ANC, SBA, and PNC across individual, household, and community-level factors considered. We found that only 8.46% of the total mothers analyzed reported at least four or more ANC visits, SBA was reported by 35.12%, and 7.91% reported the use of PNC within 48 hours of birth. When comparing these indicators to generate CoC, we found that for SBA, among those with less than four ANC visits, the majority of births were not assisted by skilled personnel (68.7%), while among those with four or more ANC visits, most births were assisted by skilled personnel (76.7%) (Table 2). Regarding PNC, a higher proportion of women and newborns received PNC within 48 hours of birth among those with four or more ANC visits (28.3%) compared to those with less than four ANC visits (6.0%). Additionally, the prevalence of different levels of CoC varied, with the majority falling into the lowest/none category (62.9%), followed by moderate (34.7%), and the highest (WHO recommended level) (2.4%). We also found noticeable differences in CoC across individual, household, and community-level factors we considered.

**Table 2:** Grouping of maternal healthcare indicators (antenatal healthcare visits, skilled birth attendance during delivery and postnatal healthcare visits) into the level of continuum of care (highest, moderate, lowest/none) in Bangladesh, SHDS, 2020 (N= 7,079)

| Delivery and postnatal healthcare visits | Antenatal healthcare visits |  |
| --- | --- | --- |
|  | Less than 4 ANC visits<br>(n=6480; 100%) | 4 or more ANC visits<br>(n=599; 100%) |
| Delivery healthcare |  |  |
| Birth assisted by skilled personnel (n=2486; 100%) | Middle (2027; 31.3%) | Highest (560; 76.7%) |
| Birth not assisted by skilled personnel (n=4593; 100%) | Lowest (4453; 68.7%) | Middle (139; 23.3%) |
| Postnatal healthcare for women and newborns |  |  |
| Received postnatal healthcare within 48 hours of birth (n=560; 100%) | Middle (390; 6.0%) | Highest (170; 28.3%) |
| Not received postnatal healthcare or either women or newborns was missed within 48 hours of birth (n=6519; 100%) | Lowest (6090; 94.0) | Middle (429; 71.7%) |
| Prevalence of levels of healthcare-seeking behaviors |  |  |
| Level of the continuum of maternal healthcare | Number | Percent (95% CI) |
| CoC (Lowest/none) | 4452 | 62.9 (60.2-65.5) |
| CoC (Moderate) | 2457 | 34.7 (32.2-37.3) |
| CoC (Highest; WHO recommended level) | 170 | 2.4 (1.9-3.1) |

**Notes:** \*Classifications reflect current World Health Organization guidelines; N=total number of women included in the study; n= number of women who used a particular service(s).

### Continuity of Care in Accessing Healthcare Services from Antenatal to Postnatal Healthcare by Pregnancy Intention at Conception

We calculated the CoC level for overall and across pregnancy intentions and presented it in Figure 1 (a-d). We found that approximately 32% of the total sample reported at least one ANC, which declined to only 2.4% upon receiving PNC. Approximately 73% of the total mothers who reported using at least one ANC dropped out before reaching at least four ANC visits. The second-highest dropout was reported when transitioning from SBA to PNC. This dropout experience for the overall sample was mirrored across pregnancy intentions. Among mothers who reported a wanted pregnancy at conception, 38.2% reported access to at least one ANC, and this figure declined to 3.0% for those accessing the highest level of PNC. Moreover, among mothers with mistimed pregnancies and unwanted pregnancies, only 1.6% and 1.1%, respectively, reported the highest level of CoC.

**Figure 1:**
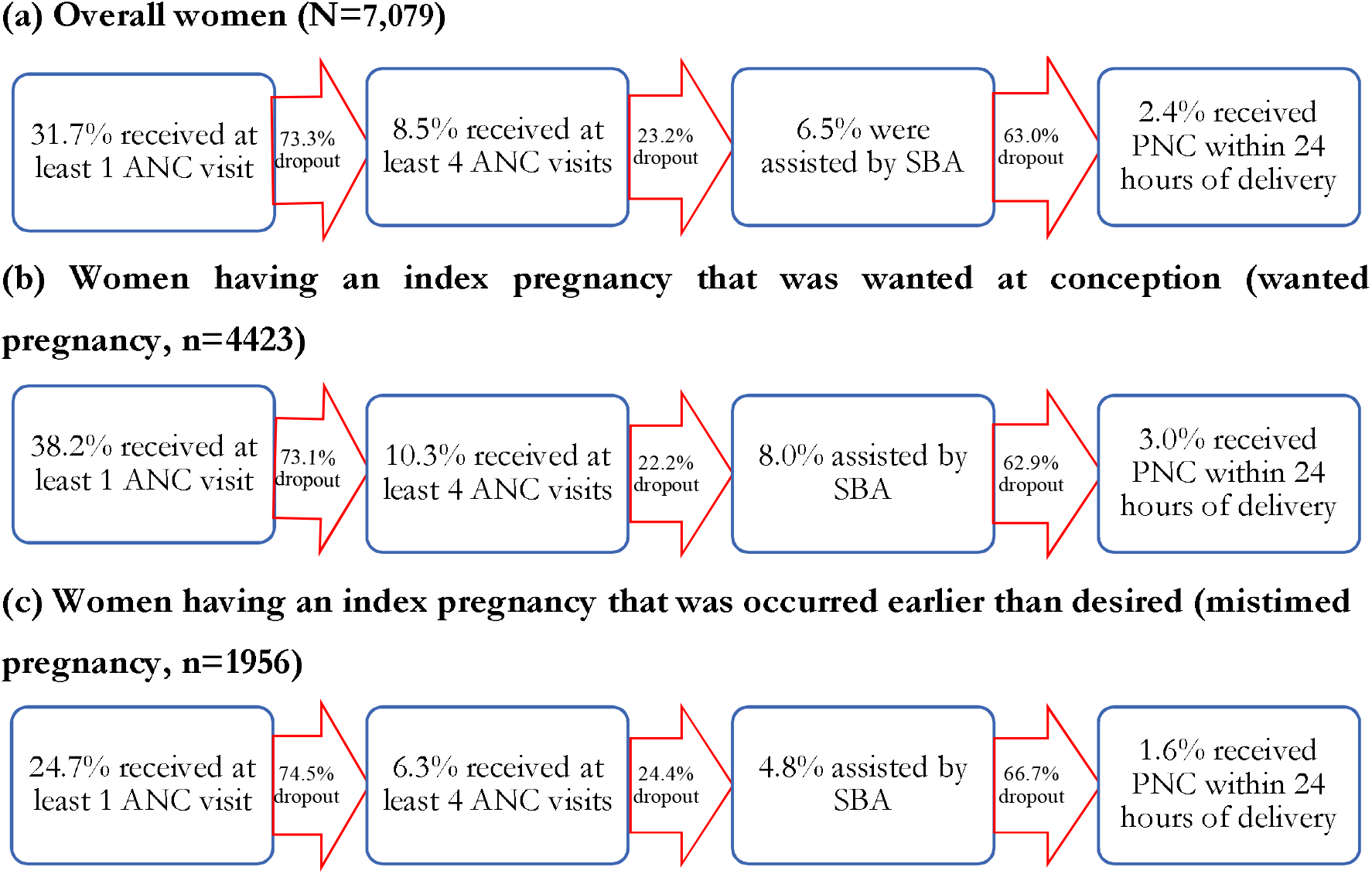
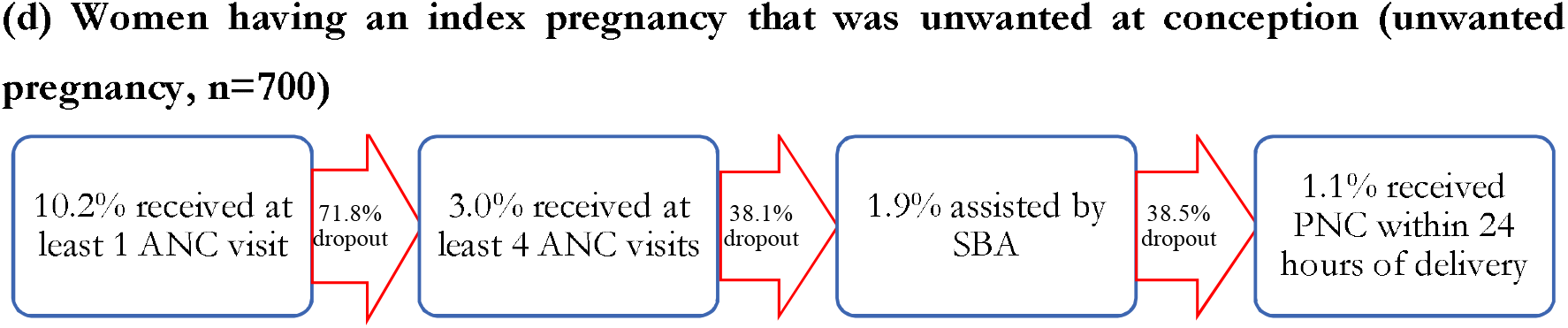
Women who drop out of the continuum of maternal healthcare through to PNC by women’s pregnancy intention at conception of their last pregnancy that ended with a live birth in Somali.

### Effects of Pregnancy Intention on Continuity of Care in Maternal Healthcare Services Use

We examined the effects of mothers’ pregnancy intention at conception on the level of CoC through unadjusted (Table 3) and adjusted (Table 4) multilevel multinomial logistic regression models. In the unadjusted model, we observed an approximately 36% lower likelihood (OR, 0.74, 95% CI, 0.62-0.90) of moderate CoC and an 81% lower likelihood (OR, 0.19, 95% CI, 0.07-0.51) of higher CoC among mothers having unwanted pregnancies compared to those with wanted pregnancies. Mothers having mistimed pregnancies had a 49% lower likelihood (OR, 0.51, 95% CI, 0.33-0.78) of achieving higher CoC compared to mothers with wanted pregnancies at conception.

**Table 3.**
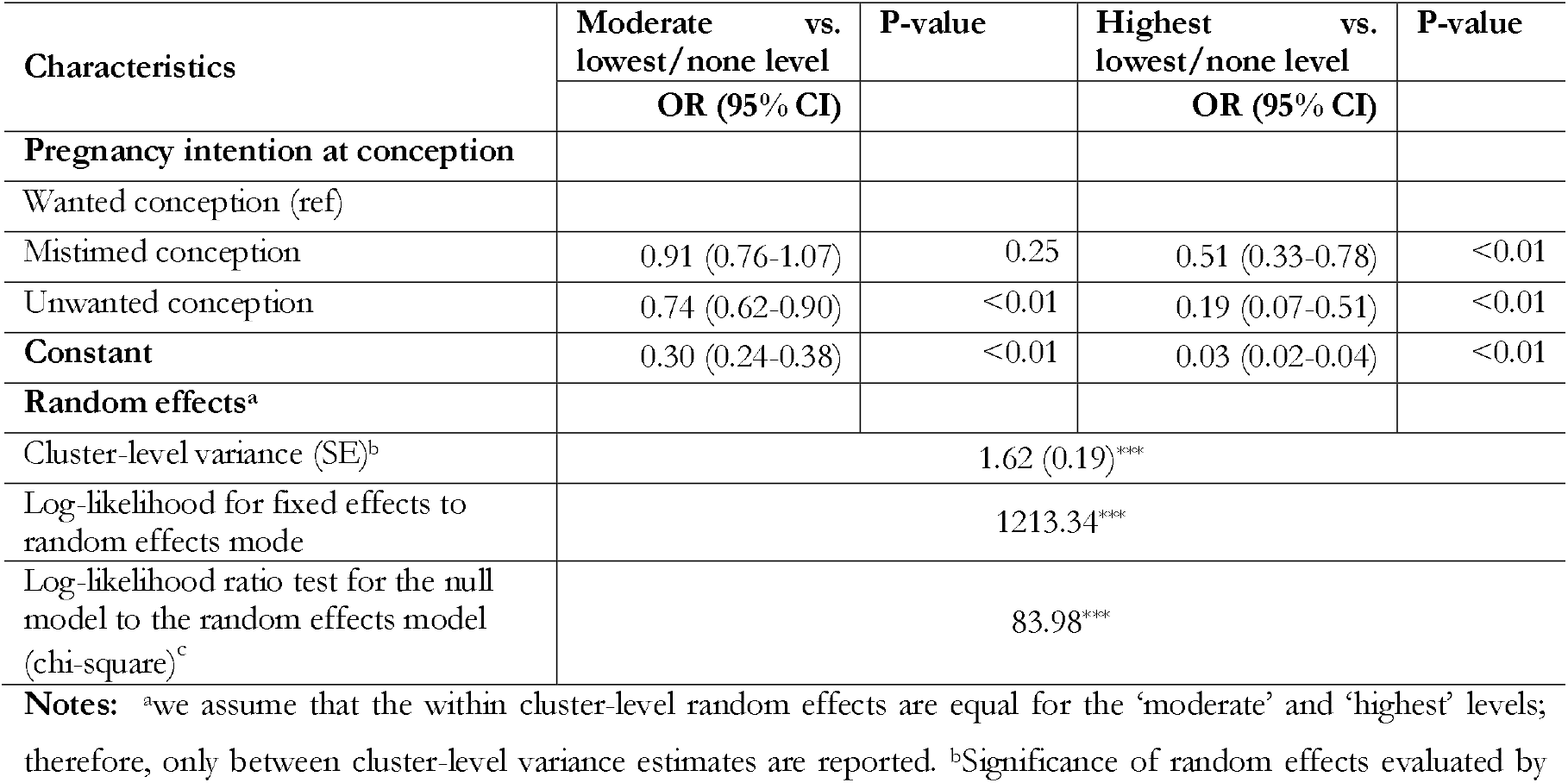
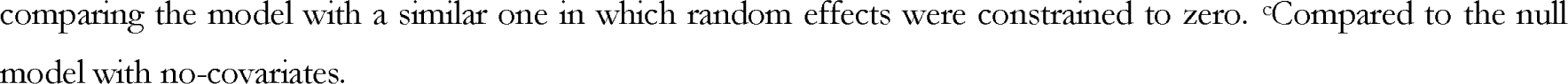
Results from multilevel multinomial logistic regression model in assessing the association between intention at conception of mother’s most recent pregnancy and level of continuum of maternal healthcare, SHDS, 2020.

**Table 4.**
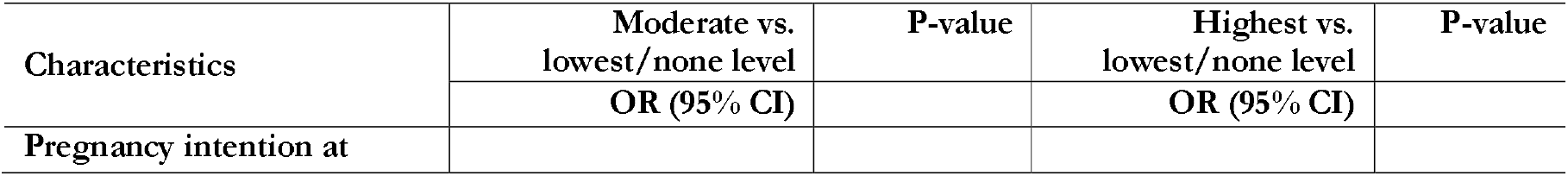
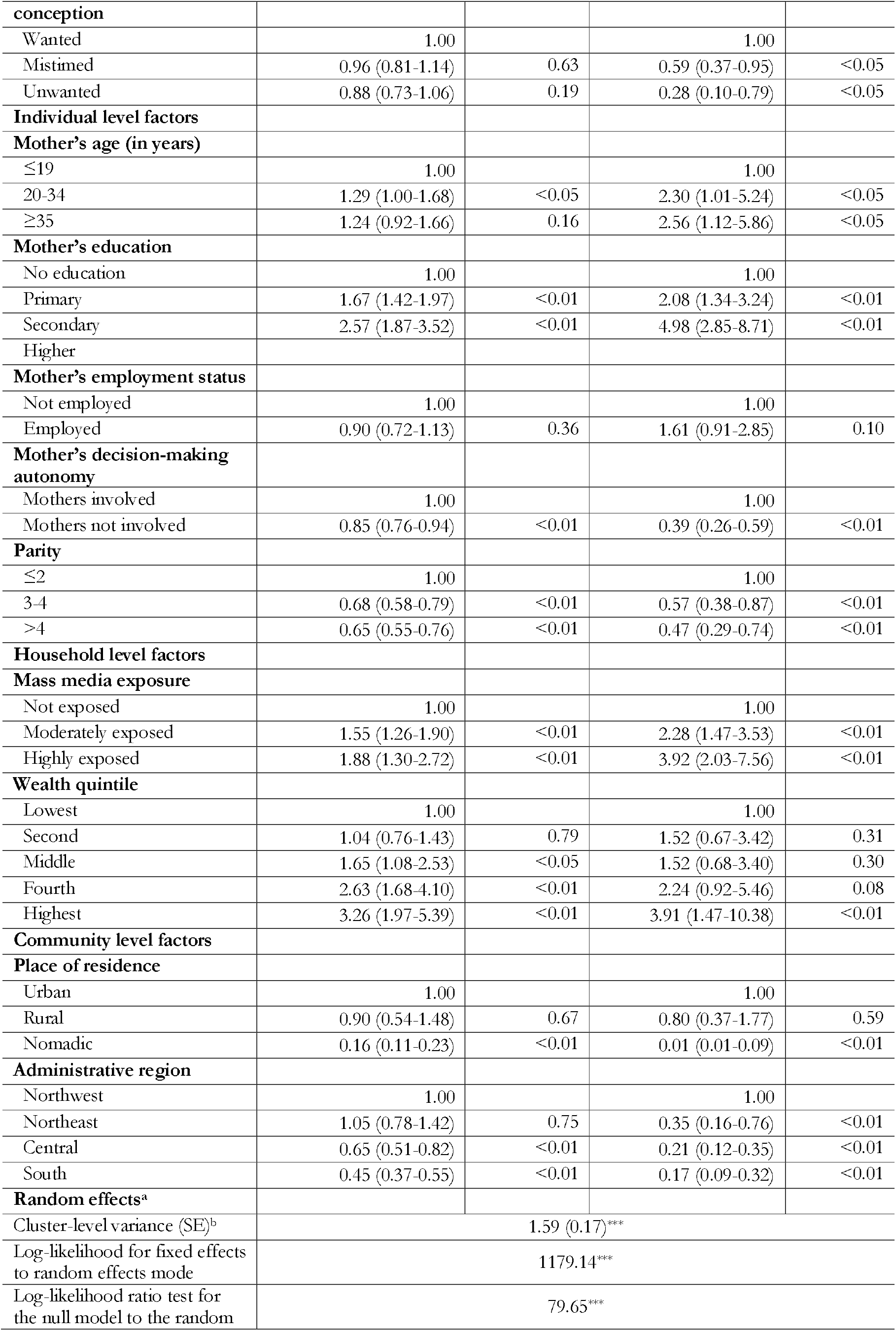
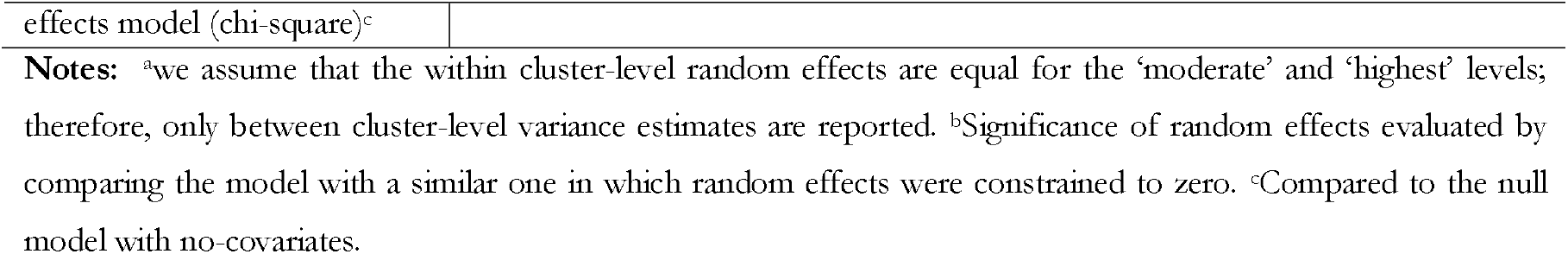
Results from multilevel multinomial logistic regression to assess the association between level of continuum of care and mother’s pregnancy intention at conception, adjusting for individual-, household-, and community-level factors, SHDS, 2020.

| Characteristics | Moderate vs.<br>lowest/none level | P-value | Highest vs.<br>lowest/none level | P-value |
| --- | --- | --- | --- | --- |
|  | OR (95% CI) |  | OR (95% CI) |  |
| Pregnancy intention at |  |  |  |  |
| **conception** |  |  |  |  |
| Wanted | 1.00 |  | 1.00 |  |
| Mistimed | 0.96 (0.81-1.14) | 0.63 | 0.59 (0.37-0.95) | <0.05 |
| Unwanted | 0.88 (0.73-1.06) | 0.19 | 0.28 (0.10-0.79) | <0.05 |
| **Individual level factors** |  |  |  |  |
| **Mother's age (in years)** |  |  |  |  |
| ≤19 | 1.00 |  | 1.00 |  |
| 20-34 | 1.29 (1.00-1.68) | <0.05 | 2.30 (1.01-5.24) | <0.05 |
| ≥35 | 1.24 (0.92-1.66) | 0.16 | 2.56 (1.12-5.86) | <0.05 |
| **Mother's education** |  |  |  |  |
| No education | 1.00 |  | 1.00 |  |
| Primary | 1.67 (1.42-1.97) | <0.01 | 2.08 (1.34-3.24) | <0.01 |
| Secondary | 2.57 (1.87-3.52) | <0.01 | 4.98 (2.85-8.71) | <0.01 |
| Higher |  |  |  |  |
| **Mother's employment status** |  |  |  |  |
| Not employed | 1.00 |  | 1.00 |  |
| Employed | 0.90 (0.72-1.13) | 0.36 | 1.61 (0.91-2.85) | 0.10 |
| **Mother's decision-making autonomy** |  |  |  |  |
| Mothers involved | 1.00 |  | 1.00 |  |
| Mothers not involved | 0.85 (0.76-0.94) | <0.01 | 0.39 (0.26-0.59) | <0.01 |
| **Parity** |  |  |  |  |
| ≤2 | 1.00 |  | 1.00 |  |
| 3-4 | 0.68 (0.58-0.79) | <0.01 | 0.57 (0.38-0.87) | <0.01 |
| >4 | 0.65 (0.55-0.76) | <0.01 | 0.47 (0.29-0.74) | <0.01 |
| **Household level factors** |  |  |  |  |
| **Mass media exposure** |  |  |  |  |
| Not exposed | 1.00 |  | 1.00 |  |
| Moderately exposed | 1.55 (1.26-1.90) | <0.01 | 2.28 (1.47-3.53) | <0.01 |
| Highly exposed | 1.88 (1.30-2.72) | <0.01 | 3.92 (2.03-7.56) | <0.01 |
| **Wealth quintile** |  |  |  |  |
| Lowest | 1.00 |  | 1.00 |  |
| Second | 1.04 (0.76-1.43) | 0.79 | 1.52 (0.67-3.42) | 0.31 |
| Middle | 1.65 (1.08-2.53) | <0.05 | 1.52 (0.68-3.40) | 0.30 |
| Fourth | 2.63 (1.68-4.10) | <0.01 | 2.24 (0.92-5.46) | 0.08 |
| Highest | 3.26 (1.97-5.39) | <0.01 | 3.91 (1.47-10.38) | <0.01 |
| **Community level factors** |  |  |  |  |
| **Place of residence** |  |  |  |  |
| Urban | 1.00 |  | 1.00 |  |
| Rural | 0.90 (0.54-1.48) | 0.67 | 0.80 (0.37-1.77) | 0.59 |
| Nomadic | 0.16 (0.11-0.23) | <0.01 | 0.01 (0.01-0.09) | <0.01 |
| **Administrative region** |  |  |  |  |
| Northwest | 1.00 |  | 1.00 |  |
| Northeast | 1.05 (0.78-1.42) | 0.75 | 0.35 (0.16-0.76) | <0.01 |
| Central | 0.65 (0.51-0.82) | <0.01 | 0.21 (0.12-0.35) | <0.01 |
| South | 0.45 (0.37-0.55) | <0.01 | 0.17 (0.09-0.32) | <0.01 |
| **Random effects<sup>a</sup>** |  |  |  |  |
| Cluster-level variance (SE)<sup>b</sup> | 1.59 (0.17)<sup>***</sup> |  |  |  |
| Log-likelihood for fixed effects to random effects mode | 1179.14<sup>***</sup> |  |  |  |
| Log-likelihood ratio test for the null model to the random | 79.65<sup>***</sup> |  |  |  |

|  |
| --- |
| effects model (chi-square)<sup>c</sup> |
| **Notes:** <sup>a</sup>we assume that the within cluster-level random effects are equal for the ‘moderate’ and ‘highest’ levels; therefore, only between cluster-level variance estimates are reported. <sup>b</sup>Significance of random effects evaluated by comparing the model with a similar one in which random effects were constrained to zero. <sup>c</sup>Compared to the null model with no-covariates. |

Once we adjusted the unadjusted model with individual, household, and community-level factors, we found that mothers with mistimed and unwanted pregnancies reported 41% (aOR, 0.59, 95% CI, 0.37-0.95) and 72% (aOR, 0.28, 95% CI, 0.10-0.79) lower likelihoods of achieving a higher level of CoC compared to mothers with wanted pregnancies.

Among the adjusted factors, mothers aged 20-34 years and ≥35 years had significantly higher odds of achieving the highest level of CoC compared to those aged ≤19 years. Mothers with primary, secondary, and higher education levels had significantly higher odds of achieving both moderate and highest levels of CoC compared to those with no education. Conversely, mothers not involved in decision-making about healthcare had significantly lower odds of achieving both moderate and highest levels of CoC compared to those involved. Regarding parity, mothers with 3-4 and >4 children had significantly lower odds of achieving both moderate and highest levels of CoC compared to those with ≤2 children. Mass media exposure and wealth quintile also showed significant associations with CoC levels, with higher exposure and wealth associated with increased odds of achieving moderate and highest levels of CoC. Living in a nomadic region was associated with a significantly lower likelihood of achieving the highest level of CoC. Additionally, mothers residing in the northeast, central, and south regions reported lower likelihoods of achieving higher and moderate levels of CoC compared to those residing in the northwest region. These factors were also found to be associated with each component of maternal healthcare services use (S1-Table 2).

## Discussion

This study aimed to investigate the effect of unintended pregnancy on the CoC in maternal healthcare services in Somalia. Our findings revealed that approximately 38% of live births in Somalia were unintended at conception. Only 2.4% of mothers reported achieving CoC, with higher rates observed among those who intended their pregnancy (3.0%) compared to those with mistimed (1.6%) or unwanted pregnancies (1.1%). Mothers with mistimed and unwanted pregnancies had significantly lower odds of achieving moderate and higher levels of CoC compared to those with intended pregnancies, even after adjusting for various socio-demographic factors. These results highlight the significant burden of unintended pregnancy in Somalia, both quantitatively and in terms of its impact on accessing maternal healthcare services and maintaining CoC. Strengthening family planning services and promoting contraceptive use are crucial steps in addressing this issue effectively.

We observed that approximately 38% of live births in Somalia are unintended at conception, a rate notably higher than the average in LMICs (8.2% of total live births) and Sub-Saharan Africa (9.1% of total live births). Limited access to family planning services, with only 8% utilizing contraception and less than 1% using modern methods, is a significant contributing factor to such higher occurrence of unintended pregnancy (1, 22, 23). Additionally, the unmet need for contraception is notably high at around 23% (24). This lack of access is compounded by inadequate sexual education and limited autonomy for women in making reproductive decisions and accessing contraception (25, 26). Cultural and societal barriers, such as early marriage and traditional gender norms, further impede access to contraception (27). Moreover, low levels of education contribute to a lack of awareness about family planning, while stigma surrounding reproductive health perpetuates misconceptions and misinformation about contraceptive methods (28, 29). These complex challenges exacerbate the prevalence of unintended pregnancies in Somalia, underscoring the critical need for comprehensive reproductive health programs and improved access to family planning services.

We found that only 2.4% of mothers in Somalia received all forms of maternal healthcare services. In comparison, the average CoC rates in sub-Saharan Africa (13.9% CoC) and South Asia (24.5% CoC) are approximately 7 to 12 times higher, highlighting Somalia’s status among countries with the lowest CoC at the global level (30). The most significant dropouts occurred between receiving at least one ANC and four or more ANC visits, as well as between SBA and PNC, reflecting the experiences of countries in sub-Saharan Africa and South Asia (30-32). Several factors may contribute to the lower prevalence of CoC and higher dropout rates in Somalia. Challenges exist at both the community and health facility levels (1, 18, 33). Limited education coverage, particularly among mothers, coupled with the absence of sexual and reproductive health education in the curriculum, may severely impact awareness about maternal healthcare services (23, 24). This contributes to lower access to CoC and affects the transition from one stage to another in CoC. Cultural and social factors, such as early marriage and traditional practices of staying home during pregnancy, also hinder access to healthcare and discourage timely medical assistance-seeking behaviors (18, 34). Additionally, poor healthcare infrastructure and inadequate facility-level efforts further exacerbate the situation (35). The ongoing conflict and instability in Somalia further disrupt healthcare delivery systems and damage infrastructure, compounding the challenges faced in accessing maternal healthcare services.

One important factor revealed in this study was the mothers’ pregnancy intention at conception, with a reduction of moderate to the highest level of CoC observed among mothers with unwanted and mistimed pregnancies, ranging from 41 to 72%. These findings can be attributed to the general challenges mentioned above along with difficulties posed by this particular type of pregnancy. Unintended pregnancies often result in additional challenges and stress for mothers and their partners, including feelings of depression and anxiety (17, 19, 36). Ensuring healthcare services in the face of such pregnancy-related challenges, amidst broader community-level challenges, may not always be feasible unless there is a strong government initiative to do so by ensuring services are available at the household level with close monitoring for continuity. However, such a focus is currently lacking in the government policies of Somalia (10, 23, 37). Additionally, unintended pregnancies may be associated with lower levels of preparedness and planning, resulting in delays in seeking healthcare and inadequate utilization of services. Such preparedness comes with disadvantages associated with mothers’ characteristics, such as limited decision-making autonomy, higher number of pregnancies, and exposure to mass media, as found in this study. These findings align with similar studies and highlight additional challenges beyond those related to pregnancy (16, 20, 21, 31). Moreover, cultural and societal stigmas surrounding unintended pregnancies may deter mothers from seeking timely medical assistance and continuing with maternal healthcare services (34, 38). Furthermore, the lack of comprehensive family planning programs and limited access to contraception exacerbate the issue, increasing the likelihood of unintended pregnancies and subsequent dropout from maternal healthcare services (23, 37).

These difficulties are even further exacerbated by partner-level challenges, where partners are often unsupportive in the case of unintended pregnancies, particularly in the event of a child’s sex being opposite to the partner’s desire or in higher-order births (15). Moreover, partners in Somalia are often overlooked as a major target group to ensure maternal healthcare services as like other LMICs (1, 16). This, along with uninformed partners—a common characteristic of unintended pregnancies—may result in partners not recognizing pregnancy symptoms or the importance of maternal health care and CoC (15). Partners might exert pressure to delay or forgo seeking medical attention due to financial concerns, fear of judgment, or cultural beliefs (15). Additionally, the lack of partner involvement in pregnancy care can leave mothers feeling alone and apprehensive about navigating the healthcare system, ultimately leading to decreased service utilization (39).

This study has several strengths and a few limitations. Notably, it is the inaugural nationally representative investigation conducted in Somalia, offering valuable insights into the current estimate of CoC and its association with maternal pregnancy intention. The study adhered to international guidelines for classifying the outcome variable, ensuring a standardized approach. Rigorous data analysis techniques, including advanced statistical methods, were employed, and a wide array of confounders were considered in the model, rendering the findings robust and informative for Somalia’s government and development partners in crafting effective policies and programs to bolster CoC. However, it is crucial to recognize the primary limitation of this study, which stems from the analysis of cross-sectional data, limiting the reported findings to correlations rather than establishing causal relationships. Additionally, the retrospective collection of survey data introduces the potential for recall bias, although any reporting errors are likely to be random. Furthermore, factors related to health facility availability and religious considerations are pertinent in understanding maternal healthcare services utilization with continuity, yet the absence of relevant data in the survey precluded their inclusion in the model. Despite these limitations, this study offers the inaugural nationally representative evidence for Somalia, serving as a foundational resource to guide evidence-based policymaking and program development aimed at enhancing maternal healthcare services utilization.

## Conclusion

The study highlights the lower utilization of CoC in Somalia, particularly in the context of unintended pregnancies. This underscores the pressing need for Somalia to address the underlying factors contributing to unintended pregnancies and lower CoC rates. To tackle these challenges effectively, a multi-faceted approach is necessary, encompassing policy interventions, improvements in healthcare infrastructure, and community-level initiatives. Strengthening family planning services, promoting contraceptive use, enhancing sexual education, and raising awareness about the importance of maternal healthcare services are crucial steps in reducing the occurrence of unintended pregnancies and mitigating their impact on CoC.

## Authors contribution

MNK, MBA and SJK developed the study concept. MNK and MBA conducted the formal analyses. SJK and MBA drafted the manuscript. MNK and SJK reviewed the first manuscript. MNK supervised all works. All the authors critically reviewed the manuscript and approved the final version of this manuscript.

## Acknowledgements

We would like to express our gratitude to the Somali Health and Demographic Survey for providing us with access to their survey data for conducting this study. Moreover, we also acknowledge to Department of Population science, Jatiya Kabi Kazi Nazrul Islam University, Bangladesh where the study was undertaken.

## Data availability

We are not authorized to share the data file with any parties. However, those interested in accessing this data can do so by submitting a research proposal to the Somalia National Data Archive authority. They can be reached by visiting the following website: https://microdata.nbs.gov.so/index.php/catalog/50

## References

1. Directorate of National Statistics, Federal Government of Somalia. The Somali Health and Demographic Survey 2020.

2. UNICEF. Monitoring the situation of children and women. Maternal mortality. 2023.

3. USAID. Yemen Health Fact Sheet. Accessed from: 01 April, 2024. https://www.usaid.gov/yemen/fact-sheets/health.

4. World Health Organization. 2018. South Sudan. Maternal Mortality. Accessed from: 01 April, 2024. https://iris.who.int/bitstream/handle/10665/136881/ccsbrief_ssd_en.pdf;sequence=1#:~:text=South%20Sudan%20has%20some%20of%20the%20worst%20health,under-five%20mortality%20rates%20are%2039.3%20and%2099.2%2C%20per.

5. UNICEF. Monitoring the situation of children and women. Child Survival. Infant mortality rate. Somalia.

6. Morrison J, Malik SMMR. Population health trends and disease profile in Somalia 1990– 2019, and projection to 2030: will the country achieve sustainable development goals 2 and 3? BMC Public Health. 2023;23(1):66.

7. Doku DT, Bhutta ZA, Neupane S. Associations of women’s empowerment with neonatal, infant and under-5 mortality in low-and/middle-income countries: meta-analysis of individual participant data from 59 countries. BMJ Global Health. 2020;5(1):e001558.

8. United Nations. Sustainable Development Goal-3. Target 3.1 Maternal mortality. Accessed from: 01 April, 2024. https://unric.org/en/sdg-3/.

9. Aden J. Maternal Health Outcomes in a Somalia Post-war Context: a PhD thesis analyzing trends towards universal health coverage. Somali Health Action Journal. 2022;2(1).

10. Mohamed AA, Bocher T, Magan MA, Omar A, Mutai O, Mohamoud SA, et al. Experiences from the field: a qualitative study exploring barriers to maternal and child health service utilization in IDP settings Somalia. International Journal of Women’s Health. 2021:1147–60.

11. Miikkulainen A, Abdirahman Mohamud I, Aqazouz M, Abdullahi Suleiman B, Sheikh Mohamud O, Ahmed Mohamed A, et al. Antenatal care utilization and its associated factors in Somalia: a cross-sectional study. BMC Pregnancy and Childbirth. 2023;23(1):581.

12. Bearak J, Popinchalk A, Ganatra B, Moller A-B, Tunçalp Ö, Beavin C, et al. Unintended pregnancy and abortion by income, region, and the legal status of abortion: estimates from a comprehensive model for 1990–2019. The Lancet Global Health. 2020;8(9):e1152–e61.

13. Aragaw FM, Amare T, Teklu RE, Tegegne BA, Alem AZ. Magnitude of unintended pregnancy and its determinants among childbearing age women in low and middle-income countries: evidence from 61 low and middle income countries. Front Reprod Health. 2023;5:1113926. Epub 20230717.

14. UNFPA. Somalia. Trusting the process: family planning and contraceptive uptake in Somalia. 2023.

15. Khan MN, Harris ML, Shifti DM, Laar AS, Loxton D. Effects of unintended pregnancy on maternal healthcare services utilization in low-and lower-middle-income countries: systematic review and meta-analysis. International journal of public health. 2019;64:743-54.

16. Khan MN, Harris ML, Oldmeadow C, Loxton D. Effect of unintended pregnancy on skilled antenatal care uptake in Bangladesh: analysis of national survey data. Archives of Public Health. 2020;78:1-13.

17. Qiu X, Zhang S, Sun X, Li H, Wang D. Unintended pregnancy and postpartum depression: A meta-analysis of cohort and case-control studies. Journal of Psychosomatic Research. 2020;138:110259.

18. Sheikh NS, Gele A. Factors influencing the motivation of maternal health workers in conflict setting of Mogadishu, Somalia. PLOS Global Public Health. 2023;3(3):e0001673.

19. Bahk J, Yun S-C, Kim Y-m, Khang Y-H. Impact of unintended pregnancy on maternal mental health: a causal analysis using follow up data of the Panel Study on Korean Children (PSKC). BMC pregnancy and childbirth. 2015;15:1–12.

20. Khan MN, Harris ML, Loxton D. Does unintended pregnancy have an impact on skilled delivery care use in Bangladesh? A nationally representative cross-sectional study using Demography and Health Survey data. Journal of Biosocial Science. 2021;53(5):773–89.

21. Khan MN, Harris ML, Loxton D. Low utilisation of postnatal care among women with unwanted pregnancy: A challenge for Bangladesh to achieve Sustainable Development Goal targets to reduce maternal and newborn deaths. Health & Social Care in the Community. 2022;30(2):e524–e36.

22. World Health Organization. High rates of unintended pregnancies linked to gaps in family planning services. 2019. Available from: https://www.who.int/news/item/25-10-2019-high-rates-of-unintended-pregnancies-linked-to-gaps-in-family-planning-services-new-who-study.

23. Yasin I, Alam MB, Khan M, Khanam SJ, Sumon K. Contraception Crisis in Somalia: Unveiling the Shocking Reality of a Nation Left Behind. Available at SSRN 4460167.

24. Alam MB, Billah MA, Yasin I, Khanam SJ, Khan MN. Unmet Contraception Need Among Married Women in Somalia: Findings from the First National Health and Demographic Survey. 2023.

25. Zhang Y, McCoy EE, Scego R, Phillips W, Godfrey E. A qualitative exploration of Somali refugee women’s experiences with family planning in the US. Journal of Immigrant and Minority Health. 2020;22:66–73.

26. Royer PA, Olson LM, Jackson B, Weber LS, Gawron L, Sanders JN, et al. “In Africa, there was no family planning. Every year you just give birth”: Family planning knowledge, attitudes, and practices among Somali and Congolese refugee women after resettlement to the United States. Qualitative health research. 2020;30(3):391.

27. Degni F, Mazengo C, Vaskilampi T, Essén B. Religious beliefs prevailing among Somali men living in Finland regarding the use of the condom by men and that of other forms of contraception by women. The European Journal of Contraception & Reproductive Health Care. 2008;13(3):298–303.

28. Davidson AS, Fabiyi C, Demissie S, Getachew H, Gilliam ML. Is LARC for everyone? A qualitative study of sociocultural perceptions of family planning and contraception among refugees in Ethiopia. Maternal and child health journal. 2017;21:1699–705.

29. Degni F, Koivusilta L, Ojanlatva A. Attitudes towards and perceptions about contraceptive use among married refugee women of Somali descent living in Finland. The European Journal of Contraception & Reproductive Health Care. 2006;11(3):190–6.

30. Singh K, Story WT, Moran AC. Assessing the continuum of care pathway for maternal health in South Asia and sub-Saharan Africa. Maternal and child health journal. 2016;20:281–9.

31. Khan MN, Harris ML, Loxton D. Assessing the effect of pregnancy intention at conception on the continuum of care in maternal healthcare services use in Bangladesh: Evidence from a nationally representative cross-sectional survey. PloS one. 2020;15(11):e0242729.

32. Bobo FT, Asante A, Woldie M, Dawson A, Hayen A. Evaluating equity across the continuum of care for maternal health services: analysis of national health surveys from 25 sub-Saharan African countries. International Journal for Equity in Health. 2023;22(1):239.

33. OCHA. United Nations Office for the Coordination of Humanitarian Affairs. Impact of Female Community Influencers in averting maternal mortality and increasing uptake of modern health care services in Gedo, South Central Somalia. 2021.

34. Francis SA, Griffith F, Leser KA. An investigation of Somali Women’s beliefs, practices, and attitudes about health, health promoting Behaviours and Cancer prevention. Health, Culture and Society. 2014;6(1):1–12.

35. World Health Organization, Somalia: building a stronger primary health care system. 2020: https://www.who.int/news-room/feature-stories/detail/somalia-building-a-strongerprimary-health-care-system.

36. Moreau C, Bonnet C, Beuzelin M, Blondel B. Pregnancy planning and acceptance and maternal psychological distress during pregnancy: results from the National Perinatal Survey, France, 2016. BMC pregnancy and childbirth. 2022;22(1):162.

37. Khan MN, Khalif IY, Rana MS, Khan MMA, Khanam SJ, Alam MB. Improving the uptake of contraception, Somalia. Bulletin of the World Health Organization. 2024;102(1):75.

38. Smith W, Turan JM, White K, Stringer KL, Helova A, Simpson T, et al. Social norms and stigma regarding unintended pregnancy and pregnancy decisions: a qualitative study of young women in Alabama. Perspectives on sexual and reproductive health. 2016;48(2):73–81.

39. Khan MN, Harris ML, Huda MN, Loxton D. A population-level data linkage study to explore the association between health facility level factors and unintended pregnancy in Bangladesh. Scientific reports. 2022;12(1):15165.

