## Supplementary files for "Effect of pregnancy intention at conception on the continuity of care in maternal healthcare services use in Somalia: Evidence from first national health and demographic survey"

**S1-Table 1.** Distribution of maternal healthcare services uptake women’s individual- household- and community level factors, SHDS, 2020 (N=7079)

| **Characteristics** | **Maternal Healthcare Services use, % (95% CI)** | | |  | **Level of continuum of care,**  **% (95% CI)** | |
| --- | --- | --- | --- | --- | --- | --- |
|  | **At least 4 ANC** | **SBA during delivery** | **PNC within 48 hours for both mothers and newborns** | **Lowest/none level** | **Moderate level** | **Highest level** |
| **Total percentages** | **8.5 (7.1-10.0)** | **35.1 (32.5-37.8)** | **7.9 (6.9-9.0)** | **62.9 (60.2-65.5)** | **34.7 (32.2-37.3)** | **2.4 (1.9-3.1)** |
| **Individual level** **factors** |  |  |  |  |  |  |
| **Pregnancy intention at conception** |  |  |  |  |  |  |
| Wanted | 10.3 (8.6-12.2) | 36.5 (33.6-39.5) | 9.3 (8.1-10.7) | 61.2 (58.2-64.1) | 35.8 (33.1-38.7) | 3.0 (2.3-3.9) |
| Mistimed | 6.3 (4.9-8.1) | 35.0 (31.2-38.9) | 6.1 (4.7-7.7) | 63.4 (59.4-67.3) | 35.0 (31.3-39.0) | 1.6 (1.1-2.3) |
| Unwanted | 3.0 (1.5-5.8) | 26.9 (22.3-32.0) | 4.1 (2.4-6.7) | 72.0 (66.7-76.7) | 26.9 (22.4-31.9) | 1.1 (0.4-3.2) |
| **Women’s age** |  |  |  |  |  |  |
| ≤19 years | 4.3 (2.4-7.6) | 33.3 (27.8-39.3) | 5.5 (3.6-8.1) | 65.5 (59.6-70.9) | 33.7 (28.3-39.5) | 0.9 (0.3-2.4) |
| 20-34 years | 9.2 (7.7-10.9) | 36.4 (33.7-39.2) | 8.1 (7.0-9.4) | 61.6 (58.7-64.3) | 35.8 (33.2-38.5) | 2.7 (2.0-2.4) |
| ≥35 years | 7.5 (5.7-9.7) | 31.5 (27.9-35.3) | 8.0 (6.4-10.2) | 66.6 (62.8-70.2) | 31.4 (27.8-35.2) | 2.0 (1.3-3.2) |
| **Women’s education** |  |  |  |  |  |  |
| No education | 6.4 (5.3-7.7) | 28.7 (26.2-31.3) | 5.5 (4.7-6.5) | 69.4 (66.7-71.9) | 29.2 (26.8-31.8) | 1.4 (1.0-2.0) |
| Primary | 11.4-21.4) | 61.3 (55.8-66.5) | 15.1 (11.9-19.0) | 36.5 (31.2-42.2) | 58.7 (53.7-63.6) | 4.8 (3.1-7.2) |
| Secondary and higher | 25.7 (18.7-34.3) | 80.3 (74.0-85.4) | 31.1 (24.9-38.1) | 17.9 (13.0-24.3) | 68.7 (62.2-74.6) | 13.3 (8.9-19.5) |
| **Women’s employment status** |  |  |  |  |  |  |
| Not employed | 8.1 (6.8-9.7) | 34.5 (31.9-37.3) | 7.4 (6.5-8.5) | 63.6 (60.8-66.2) | 34.2 (31.7-36.8) | 2.2 (1.7-2.9) |
| Employed | 13.4 (8.8-19.7) | 44.3 (38.4-50.4) | 15.5 (11.4-20.5) | 51.9 (45.6-58.2) | 43.2 (36.9-49.7) | 4.9 (2.9-8.2) |
| **Decision-making about women's healthcare** |  |  |  |  |  |  |
| Women involved | 11.2 (9.1-13.6) | 40.8 (37.5-44.2) | 10.4 (8.9-12.0) | 57.1 (53.6-60.5) | 39.2 (36.2-42.3) | 3.7 (2.9-4.9) |
| Women not involved | 5.2 (4.1-6.7) | 28.4 (25.2-31.8) | 5.0 (4.0-6.1) | 69.8 (66.3-73.1) | 29.4 (26.2-32.9) | 0.8 (0.5-1.4) |
| **Parity** |  |  |  |  |  |  |
| ≤2 | 8.9 (7.0-11.3) | 39.5 (35.6-43.5) | 10.6 (8.8-12.7) | 59.1 (55.1-63.0) | 37.4 (33.7-41.3) | 3.5 (2.4-5.0) |
| 3-4 | 8.7 (6.4-11.6) | 35.3 (31.7-39.0) | 7.2 (5.6-9.2) | 62.8 (59.0-66.4) | 35.1 (31.8-38.6) | 2.1 (1.4-3.0) |
| >4 | 8.0 (6.5-9.8) | 32.2 (29.4-35.2) | 6.6 (5.5-8.0) | 65.4 (62.4-68.3) | 32.7 (29.9-35.7) | 2.0 (1.4-2.6) |
| **Household level** **factors** |  |  |  |  |  |  |
| **Mass media exposure** |  |  |  |  |  |  |
| Not exposed | 6.3 (5.2-7.7) | 30.0 (27.5-32.6) | 5.4 (4.6-6.3) | 68.2 (65.6-70.7) | 30.5 (28.1-33.0) | 1.3 (1.0-1.8) |
| Moderately exposed | 18.7 (15.3-22.7) | 57.7 (52.1-63.2) | 18.9 (15.8-22.5) | 39.1 (33.9-44.6) | 53.5 (48.8-58.2) | 7.4 (5.3-10.1) |
| Highly exposed | 19.1 (12.0-29.1) | 69.9 (60.0-78.3) | 25.3 (16.3-37.0) | 29.1 (20.8-39.1) | 62.6 (52.8-71.5) | 8.3 (4.7-14.3) |
| **Wealth quintile** |  |  |  |  |  |  |
| Lowest | 1.6 (0.8-3.2) | 10.6 (8.2-13.7) | 2.7 (1.8-4.2) | 88.5 (85.5-91.0) | 10.9 (8.6-13.7) | 0.5 (0.2-1.6) |
| Second | 3.6 (2.1-6.0) | 14.6 (11.6-18.1) | 2.8 (1.6-4.7) | 83.5 (79.9-86.6) | 15.7 (12.9-19.0) | 0.9 (0.3-2.4) |
| Middle | 8.8 (6.5-11.9) | 37.3 (33.9-40.8) | 6.0 (4.4-8.1) | 60.5 (56.9-64.0) | 37.3 (33.9-40.9) | 2.2 (1.2-3.8) |
| Fourth | 11.8 (9.4-14.8) | 54.3 (50.2-58.3) | 9.5 (7.6-11.9) | 42.7 (38.5-46.9) | 55.4 (51.3-59.5) | 2.0 (1.3-2.9) |
| Highest | 19.8 (16.5-23.6) | 69.9 (65.3-74.2) | 21.9 (18.6-25.5) | 27.9 (23.8-32.4) | 64.2 (59.8-68.4) | 7.9 (5.8-10.7) |
| **Community level** **factors** |  |  |  |  |  |  |
| **Place of residence** |  |  |  |  |  |  |
| Urban | 14.2 (11.9-16.9) | 57.1 (53.5-60.6) | 14.1 (12.3-16.2) | 39.7 (36.3-43.2) | 55.8 (52.6-59.0) | 4.5 (3.3-6.0) |
| Rural | 10.5 (7.2-15.0) | 39.3 (32.8-46.2) | 7.8 (5.8-10.4) | 58.7 (51.4-65.6) | 38.6 (32.2 (45.4) | 2.8 (1.7-4.3) |
| Nomadic | 0.9 (0.4-2.3) | 9.4 (7.5-11.7) | 1.8 (1.2-2.7) | 89.8 (87.6-91.7) | 10.2 (8.3-12.4) | 0.02 (0.01-0.2) |
| **Administrative region** |  |  |  |  |  |  |
| Northwest | 13.0 (10.6-15.8) | 33.1 (29.1-37.4) | 9.8 (8.2-11.7) | 64.3 (59.9-68.4) | 31.8 (28.0-35.8) | 4.0 (3.0-5.2) |
| Northeast | 5.0 (3.1-7.8) | 41.4 (36.9-46.1) | 8.0 (6.0-10.6) | 58.4 (53.7-62.9) | 39.9 (35.8-44.2) | 1.7 (0.9-3.4) |
| Central | 8.0 (5.9-10.8) | 35.8 (30.3-41.7) | 6.1 (4.7-8.0) | 60.9 (54.8-66.8) | 37.5 (31.8-43.5) | 1.6 (0.9-2.7) |
| South | 3.0 (2.2-4.0) | 17.8 (12.2-25.3) | 5.0 (3.2-7.8) | 80.9 (73.7-86.4) | 18.4 (13.1-25.3) | 0.7 (0.4-1.5) |

**Note:** Presented as row percentages.

**S1-Table 2.** Multilevel binary logistic regression model in assessing the relationships of each indicator of healthcare services use across women’s individual, household and community level factors in Somalia, SHDS, 2020 (N=7,079)

|  | **Independent component of maternal healthcare services use** | | |
| --- | --- | --- | --- |
|  | **At least 4 ANC** | **SBA during delivery** | **PNC within 48 hours for both mothers and newborns** |
| **Individual level** **factors** |  |  |  |
| **Pregnancy intention at conception** |  |  |  |
| Wanted | 1.00 | 1.00 | 1.00 |
| Mistimed | 0.70 (0.57-0.87)** | 0.96 (0.84-1.10) | 0.61 (0.49-0.76)** |
| Unwanted | 0.33 (0.19-0.55)** | 0.90 (0.72-1.12) | 0.46 (0.30-0.72)** |
| **Women’s age** |  |  |  |
| ≤19 years | 1.00 | 1.00 | 1.00 |
| 20-34 years | 1.60 (1.02-2.51)* | 1.34 (1.05-1.71)* | 1.49 (1.01-2.20)* |
| ≥35 years | 1.41 (0.85-2.36) | 1.35 (1.01-1.80)* | 2.01 (1.27-3.19)** |
| **Women’s education** |  |  |  |
| No education | 1.00 | 1.00 | 1.00 |
| Primary | 1.24 (0.98-1.58) | 1.69 (1.43-1.99)** | 1.41 (1.11-1.78)** |
| Secondary | 1.79 (1.30-2.46)** | 2.71 (2.02-3.65)** | 2.28 (1.70-3.06)** |
| Higher |  |  |  |
| **Women’s employment status** |  |  |  |
| Not employed | 1.00 | 1.00 | 1.00 |
| Employed | 1.12 (0.80-1.57) | 0.89 (0.70-1.12) | 1.45 (1.06-1.98)* |
| **Decision-making about women's healthcare** |  |  |  |
| Women involved | 1.00 | 1.00 | 1.00 |
| Women not involved | 0.69 (0.57-0.84)** | 0.82 (0.73-0.92)** | 0.60 (0.50-0.73)** |
| **Parity** |  |  |  |
| ≤2 | 1.00 | 1.00 | 1.00 |
| 3-4 | 0.98 (0.76-1.26) | 0.63 (0.54-0.75)** | 0.56 (0.44-0.72)** |
| >4 | 0.95 (0.74-1.22) | 0.60 (0.50-0.70)** | 0.54 (0.42-0.69)** |
| **Household level** **factors** |  |  |  |
| **Mass media exposure** |  |  |  |
| Not exposed | 1.00 | 1.00 | 1.00 |
| Moderately exposed | 1.54 (1.24-1.92)** | 1.53 (1.30-1.80)** | 1.79 (1.44-2.22)** |
| Highly exposed | 1.73 (1.08-2.77)* | 2.01 (1.32-3.07)** | 2.64 (1.73-4.03)** |
| **Wealth quintile** |  |  |  |
| Lowest | 1.00 | 1.00 | 1.00 |
| Second | 1.16 (0.71-1.88) | 1.05 (0.82-1.34) | 1.07 (0.67-1.71) |
| Middle | 1.24 (0.76-2.03) | 1.79 (1.38-2.33)** | 1.32 (0.80-2.18) |
| Fourth | 1.49 (0.90-2.44) | 2.82 (2.15-3.70)** | 2.07 (1.27-3.40)** |
| Highest | 1.67 (1.01-2.76)* | 3.56 (2.68-4.74)** | 2.78 (1.69-4.58)** |
| **Community level** **factors** |  |  |  |
| **Place of residence** |  |  |  |
| Urban | 1.00 | 1.00 | 1.00 |
| Rural | 1.10 (0.83-1.45) | 0.88 (0.72-1.09) | 0.80 (0.60-1.06)) |
| Nomadic | 0.07 (0.04-0.14)** | 0.17 (0.13-0.22)** | 0.26 (0.16-0.42)** |
| **Administrative region** |  |  |  |
| Northwest | 1.00 | 1.00 | 1.00 |
| Northeast | 0.31 (0.23-0.43)** | 1.15 (0.94-1.41) | 0.57 (0.42-0.77)** |
| Central | 0.31 (0.24-0.40)** | 0.65 (0.55-0.80)** | 0.44 (0.34-0.57)** |
| South | 0.20 (0.14-0.29)** | 0.49 (0.40-0.61)** | 0.82 (0.61-1.10) |
| **Summary statistics** |  |  |  |
| Constants | 0.14 (0.07-0.28)** | 0.64 (0.44-0.92)* | 0.11 (0.06-0.21)** |
| ICC | 4.07 | 8.32 | 3.33 |
| AIC | 3420.39 | 7257.93 | 3526.05 |
| BIC | 3578.89 | 7416.42 | 3684.54 |

**Notes:** ICC=Intra-class Correlation Coefficient. AIC=Akaike's Information Criterion. BIC=Bayesian Information Criterion. aOR: adjusted odds ratio. 95% CI: 95% Confidence Intervals. **p<0.01, *p<0.05.
